# Age- and sex-specific differences in immune responses to BNT162b2 COVID-19 and live-attenuated influenza vaccines in UK adolescents

**DOI:** 10.1101/2023.07.24.23293091

**Authors:** Cecilia Jay, Emily Adland, Anna Csala, Nicholas Lim, Stephanie Longet, Ane Ogbe, Consortium PITCH, Jeremy Ratcliff, Oliver Sampson, Craig P Thompson, Lance Turtle, Eleanor Barnes, Susanna Dunachie, Paul Klenerman, Miles Carroll, Philip Goulder

## Abstract

Key to understanding COVID-19 correlates of protection is assessing vaccine-induced immunity in different demographic groups. Sex- and age-specific immune differences have a wide impact on outcomes from infections and immunisations. Typically, adult females make stronger immune responses and have better disease outcomes but suffer more adverse events following vaccination and are more prone to autoimmune disease. To understand better the mechanisms underlying these differences in vaccine responses, we studied immune responses to two doses of BNT162b2 Pfizer COVID-19 vaccine in an adolescent cohort (n=34, ages 12-16), an age group previously shown to make significantly greater immune responses to the same vaccine compared to young adults. At the same time, we were able to evaluate immune responses to the co-administered live attenuated influenza vaccine, which has been shown to induce stronger immune responses in adult females. Blood samples from 34 adolescents taken pre- and post-vaccination with COVID-19 and influenza vaccines were assayed for SARS-CoV-2-specific IgG and neutralising antibodies, and cellular immunity specific for SARS-CoV-2 and endemic betacoronaviruses. IgG targeting influenza lineages contained in the influenza vaccine was also assessed. As previously demonstrated, total IgG responses to SARS-CoV-2 Spike antigens were significantly higher among vaccinated adolescents compared to adults (aged 32-52) who received the BNT162b2 vaccine (comparing infection-naïve, 49,696 vs 33,339; p=0.03; comparing SARS-CoV-2 previously-infected, 743,691 vs 269,985; p<0.0001) by MSD v-plex assay. However, unexpectedly, antibody responses to BNT162b2 and the live-attenuated influenza vaccine were not higher among female adolescents compared to males; among infection-naïve adolescents, antibody responses to BNT162b2 were higher in males than females (62,270 vs 36,951 p=0.008). No sex difference was identified in vaccinated adults. These unexpected findings may result from the introduction of novel mRNA vaccination platforms, generating patterns of immunity divergent from established trends, and providing new insights into what might be protective following COVID-19 vaccination.

## Introduction

The BNT162b2 Pfizer-BioNTech COVID-19 vaccine was authorised for 12-15 year olds in June 2021 in the United Kingdom as a 30μg one-dose regimen by the Medicines and Healthcare Products Regulatory Agency.[1] This was extended to a two-dose regimen in early 2022.[2] In the United Kingdom, the first dose of BNT162b2 was administered in adolescents alongside the AstraZeneca intranasal seasonal live-attenuated influenza vaccine (LAIV) FluenzTetra, therefore presenting a unique opportunity to study vaccine-induced immunity in this age group.

Older age is a primary risk factor for severe COVID-19, perhaps due to the reduced immune capacity with age driven by persistent inflammation and cellular dysfunction.[3] The death rate of COVID-19 is 0.66% overall, increasing to 7.8% in over 80s.[4] The majority of young people experience mild COVID-19; severe disease and multisystem inflammatory syndrome only occurs in a minority of paediatric patients.[5] Adolescents and children display rapid and adaptable immune responses that may contribute to improved resolution of infections, such as abundant IgM memory B-cells, broad and rapidly produced natural antibodies, and lower inflammatory cytokine responses.[6,7] Differential COVID-19 outcomes between adults and children may also be influenced by pre-existing immune responses to endemic coronaviruses that might circulate at higher levels in children.[6] Notably, adolescents between 12 and 15 years of age generate 1.76-fold higher IgG responses to BNT162b2 than 16-25 year olds, indicating either potential age-related changes in immune response even during adolescence, or an increase in cross-reactivity with endemic coronaviruses that enhances vaccine responsiveness and declines with age.[8] Finally, older individuals are more likely to have immunodeficiencies or chronic diseases which increase their risk of severe COVID-19.

In addition to age, understanding the role of sex in vaccine response is crucial for developing more effective vaccines. Adult females aged 18-49 have been shown to generate twofold greater antibody responses to trivalent influenza vaccines,[9] and adult females are more at risk for serious adverse events (SAEs) after vaccination, including after the ChAdOx1 Oxford-AstraZeneca COVID-19 vaccine. [10–12] In one study, females given half-dose influenza vaccine made marginally stronger antibody responses compared to age-matched males who received full-dose vaccine.[9] Greater vaccine-induced immune responses in females could potentially facilitate reduced dosing regimens for females, which may minimise incidence of SAEs, improve vaccine uptake, and improve vaccine supply. However, young males experience more vaccine-induced myocarditis after BNT162b2, suggesting that immune responses to mRNA vaccines may be differentially influenced by sex.[13,14] Adolescents undergoing puberty face significant changes in levels of sex hormones such as testosterone and oestrogen, which are known to modulate immunity to SARS-CoV-2 and influenza.[15,16]

To explore sex- and age-specific differences in humoral and cellular immunity to BNT162b2, we studied adolescent and adult cohorts in the United Kingdom who received this vaccine. Data collected from adolescents in this study were compared to the Protective Immunity from T-cells in Healthcare Workers (PITCH) cohort of vaccinated healthcare workers (HCWs) aged 32-52, who received two doses of BNT162b2 and also represented a mixture of previously-infected and infection-naïve individuals.[17] We explored age-specific effects on immunity within the adolescent cohort as well as between adolescents and adults. Furthermore, we examined whether sex-specific immune effects were evident. As not all adolescents also received the LAIV, we were also able to assess whether co-administration of LAIV appeared to influence the magnitude of response to BNT162b2. Furthermore, many studies of adolescent responses to BNT162b2 have used prior SARS-CoV-2 infection as an exclusion criterion[18,19]. Here, we enrolled both SARS-CoV-2 infection-naïve and previously-infected adolescents to understand the role of prior or ongoing infection in vaccine response.

## Results

### Cohort description

In November and December of 2021, 34 adolescents aged between 12 and 16 were recruited into the study through their enrolment at schools in Oxford, UK (Fig. 1A). All 34 individuals received the BNT162b2 vaccine; 26 (76%) also received the LAIV on the same day as the first dose of BNT162b2. 47% of individuals (n=16) were female, and the median age was 14.1 years (12.2-16) (Fig. 1B). 33 individuals were Caucasian, one individual was Asian. No individuals were on any regular medication.

**Figure 1:**
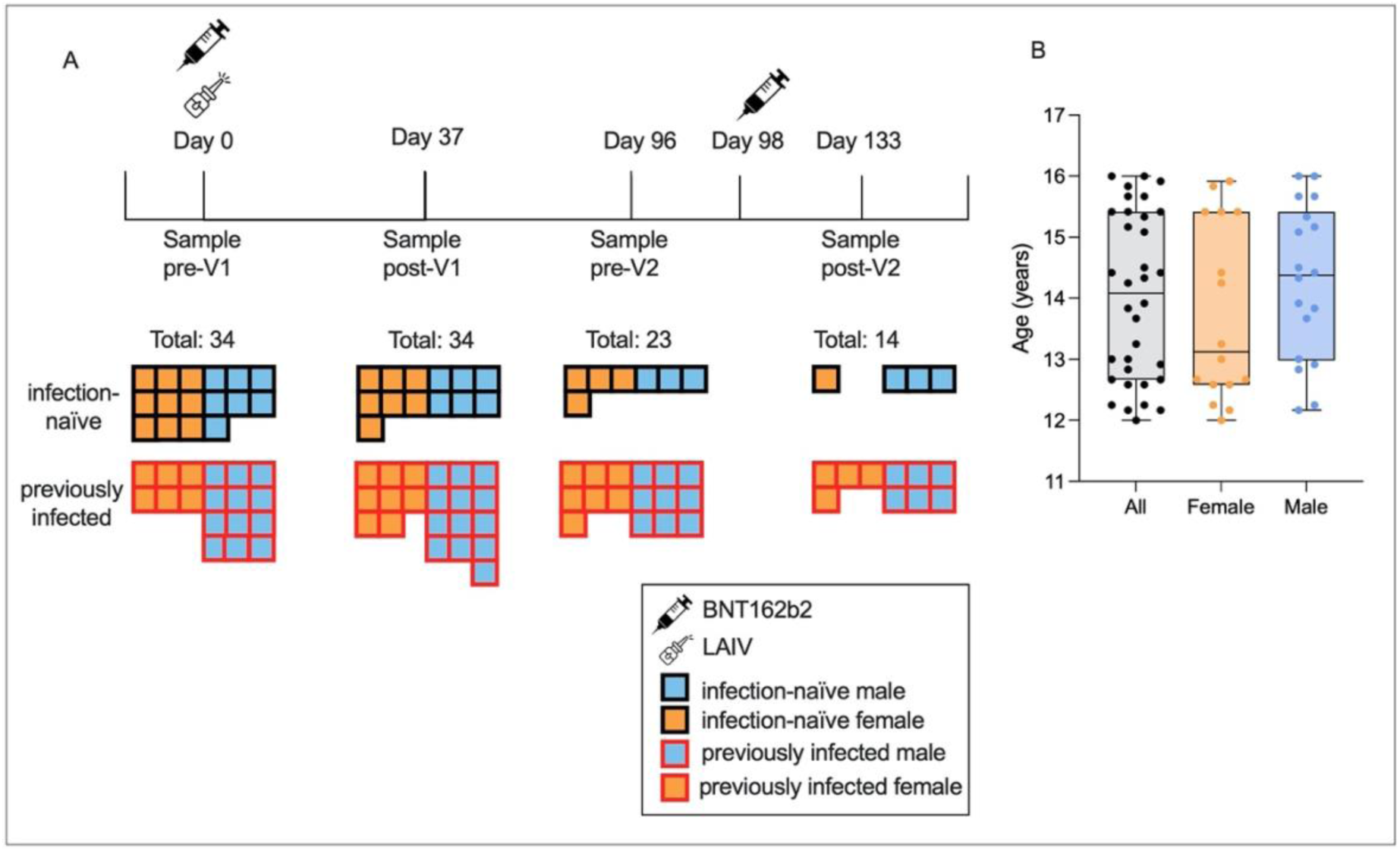
Characteristics of the study cohort. 34 adolescents were enrolled and provided consent, of which 18 were seropositive for S or N pre-V1. Samples were taken pre-V1 on the day of vaccination, a mean of 37 days post-V1, mean 2 days pre-V2, and mean 35 days post-V2 (**A**). The median age was 13 years 1 month for females (orange) and 14 years and 5 months for males (blue) (**B**).

The adult cohort to which adolescent data was compared was the PITCH cohort of vaccinated HCWs.[20,21] This cohort consisted of 589 adults aged 32-52 who had received two doses of BNT162b2 28 days apart. Here, IgG data from 170 adults and neutralising antibody (nAb) data from 10 adults was used to compare to data from adolescents.

### Humoral immune responses to BNT162b2 vaccination

To evaluate the immunogenicity of the BNT162b2 vaccine among adolescents, we first characterised humoral responses using MSD-platform immunoassays to measure quantitatively the total immunoglobulin G (IgG) response to SARS-CoV-2 Spike (S), the receptor-binding domain (RBD) of S, and SARS-CoV-2-N (Fig. 2A). Both infection-naïve and previously-infected adolescents made significantly greater IgG responses to S post-V1 than pre-V1 (61 vs 49,696, x803, p=0.0005 and 13,409 vs 788,568, x55, p<0.0001, respectively, Wilcoxon signed-rank test) and greater anti-RBD IgG responses (263 vs 16,861, x64, p=0.0005 and 6,556 vs 351,068 x53, p<0.0001, respectively, Wilcoxon signed-rank test) (Fig. 2A). Anti-S and RBD IgG responses increased post-V2 in all groups, but only anti-RBD IgG increased significantly and only in previously-infected individuals (90,067 vs 318,687, x3.5, p=0.008, Wilcoxon signed-rank test). Notably, two doses of BNT162b2 in infection-naïve individuals gave similar levels of IgG to one dose of vaccine in previously-infected individuals.

**Figure 2:**
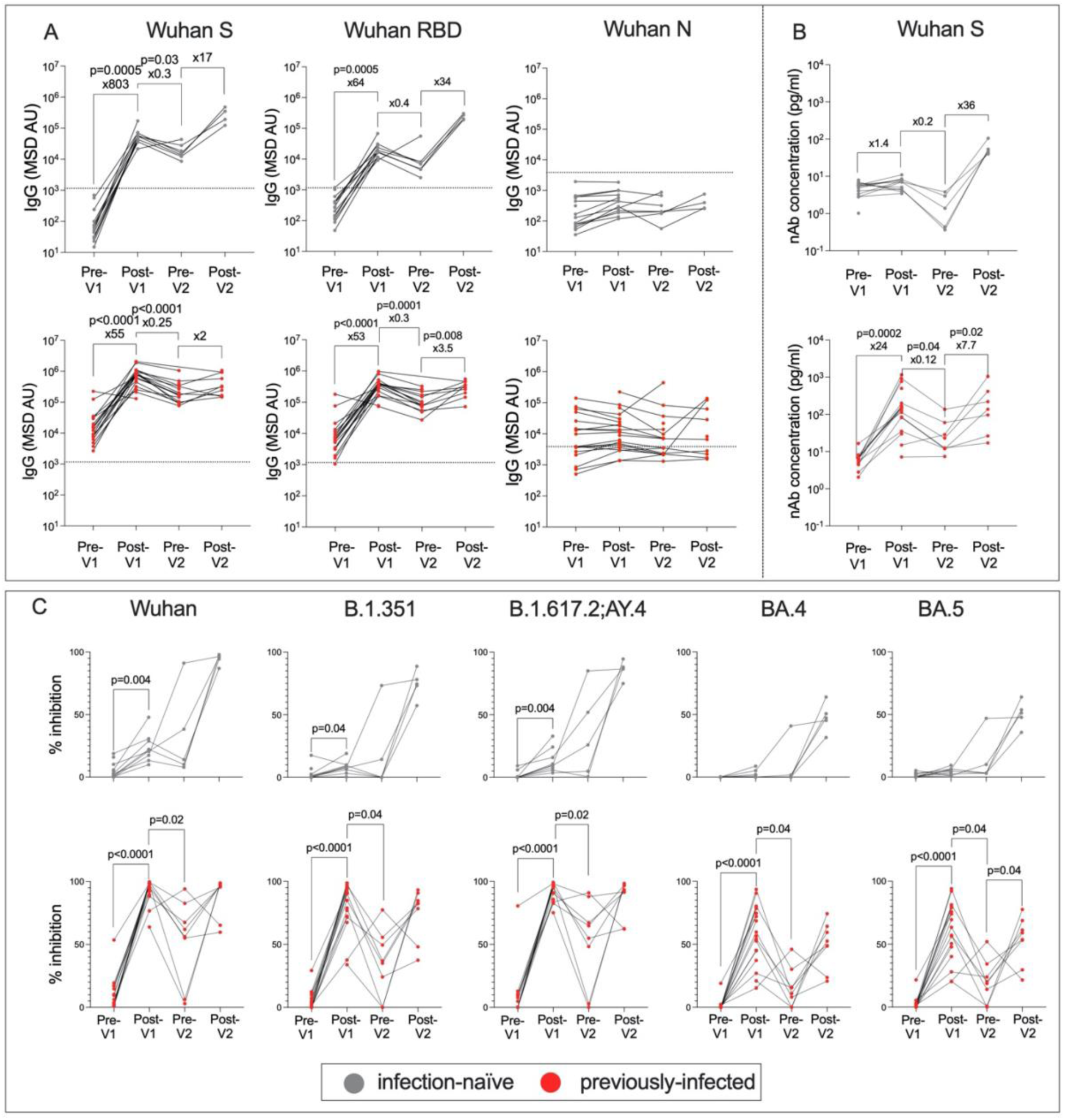
Humoral responses following first and second doses of BNT162b2 in previously-infected and infection-naive adolescents. Anti-S, RBD and N IgG in infection-naive (grey) and previously-infected (red) adolescents (**A**). Thresholds for IgG positivity were taken from previous literature.[26] nAbs targeting S in infection-naïve (grey) and previously-infected (red) adolescents using MSD ACE2-Spike binding inhibition assays (**B**). Percent inhibition of SARS-CoV-2 S-ACE2 binding as measured by MSD ACE2 inhibition assay in infection-naive (grey) and previously-infected (red) adolescents targeting common SARS-CoV-2 lineages: Wuhan, B.1.351 (Beta), B.1.617.2/AY.4 (Delta), BA.4 and BA.5 (Omicron) (**C**). P-values from Wilcoxon signed-rank tests. Fold change refers to the difference in total group medians.

Since nAbs as well as total IgG are reported to be a correlate of protective immunity against symptomatic COVID-19,[22] we next assessed a surrogate of nAb activity using the MSD-platform ACE2 inhibition assay,[23] which is well correlated with live virus neutralisation assays.[20,21,24,25] In contrast to IgG responses, only previously-infected individuals generated increased nAb responses following the first dose of vaccine (6 vs 149, x24, p=0.0002, Wilcoxon signed-rank test) (Fig. 2B), and fold change in nAb response to S and RBD was higher in previously-infected individuals post-V1 compared to infection-naïve individuals (1.1 vs 28, p<0.0001 and 1.3 vs 23, p=0.0002, respectively, Mann-Whitney tests) (Supplementary Fig. 1AB). After two doses of BNT162b2, infection-naïve individuals reached similar nAb titres to previously-infected individuals after one dose, supporting the idea that two doses of vaccine are required for a robust neutralising response in infection-naïve individuals.

To determine how breadth of nAb response to SARS-CoV-2 variants is impacted by vaccination and prior infection, the MSD-platform ACE2 inhibition assay was carried out against the common variants of SARS-CoV-2 in both infection-naïve and previously-infected individuals (Fig. 2C and Supp. Fig. 2). Notably, previously-infected individuals made broad nAb responses against all studied variants following the first dose, whereas high-titre nAb responses against these variants were only observed following the second dose in infection-naive individuals.

### Cellular immune responses to BNT162b2 vaccination

We next characterised the cellular immune response in adolescents following first and second doses of BNT162b2. Proliferation assays measuring the dilution of CellTrace Violet (CTV) stain (Thermo Fisher Scientific, USA) following stimulation of peripheral blood mononuclear cells (PBMCs) with pools of SARS-CoV-2 peptides spanning the S1 and S2 regions of S, the membrane protein (M) and N, and the S2 region of endemic human coronaviruses (HCoVs) HCoV-HKU1 and HCoV-OC43 were performed at pre-V1, post-V1, pre-V2 and post-V2 timepoints (Fig. 3, Supp. Fig. 3, Supp. File 1). In contrast to humoral responses to BNT162b2, SARS-CoV-2-specific CD4+ T-cell proliferative responses were similar in infection-naïve compared to previously-infected individuals after a single dose of vaccine. The T-cell response in previously-infected individuals increased following vaccination, but not significantly, suggesting stable cellular immunity over time.

**Figure 3:**
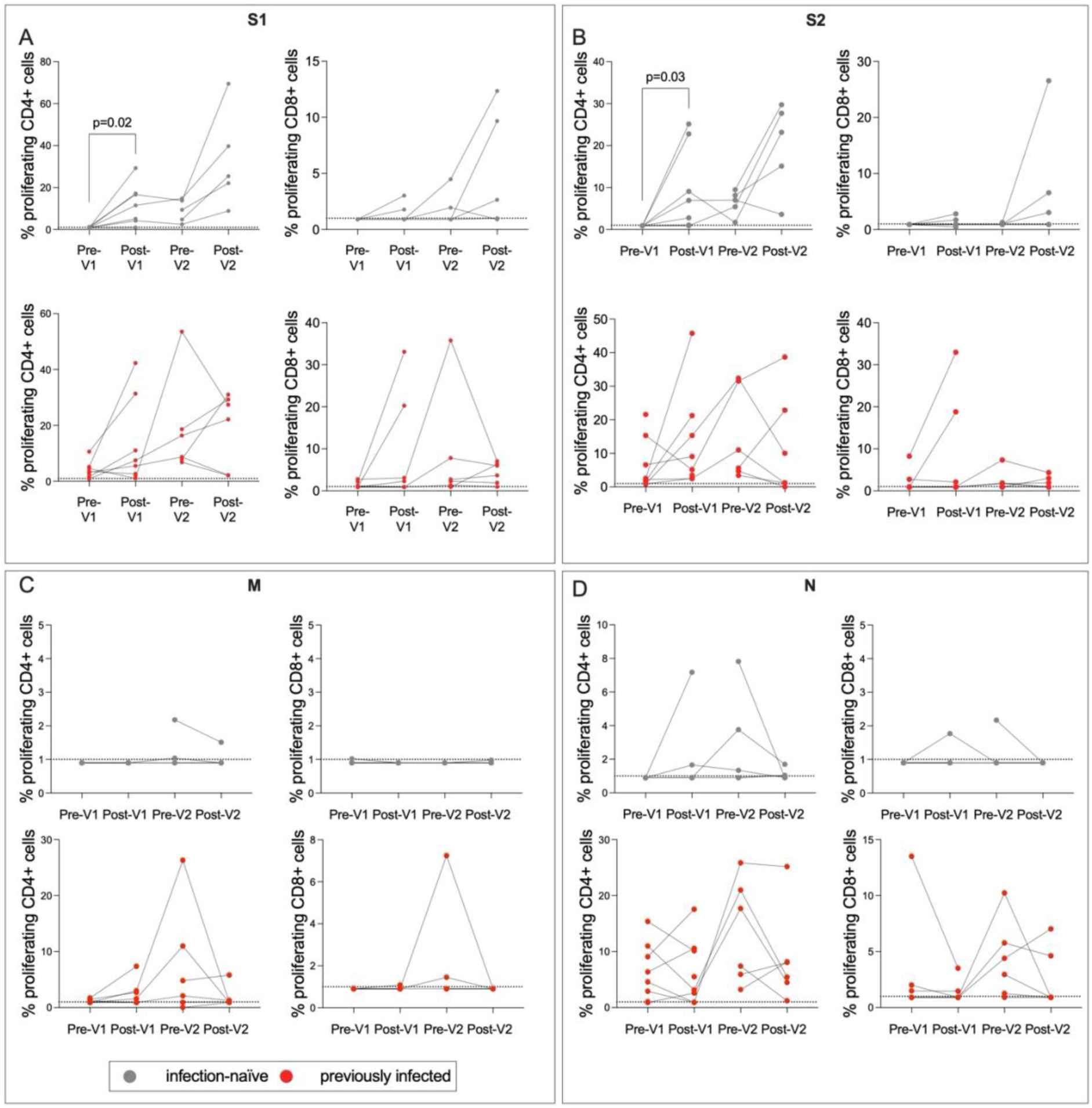
T-cell responses to SARS-CoV-2 S are boosted post-V1 and post-V2. CellTrace Violet stains were used to assess proliferating CD4+ and CD8+ T-cells targeting the S1 region of S (A), the S2 region of S (B), M (C) and N (D) in infection-naive (grey) and previously-infected (red) individuals (A). Data shows proliferating cells as a percentage of parent populations with DMSO background values subtracted. Thresholds for positivity were set at 1 as determined by previous literature.^49^ P-values from Wilcoxon signed-rank tests. Fold change refers to the difference in total group medians. Values below 1% were given nominal values of 0.9%.

Although T-cell responses to HCoV-OC43 S2 and HCoV-HKU1 S2 were identified in several individuals, particularly previously-infected individuals, there was no significant impact of BNT162b2 vaccination on the magnitude of T-cell responses.

### Higher magnitude antibody responses to BNT162b2 in adolescents versus adults

The role of age in immune response to vaccination was of particular interest in this study. To determine whether the responses observed in adolescents to the BNT162b2 vaccine were stronger than those observed in adults, as previously shown,[8] we compared the adolescent data to humoral responses in adults (32-52 years) from the PITCH cohort 28 days after the first dose of BNT162b2 (Fig. 4).[26,27] PITCH is a consortium of universities and UK Health Security Agency (UK HSA) with the aim of characterising infection-acquired and vaccine-induced immunity to SARS-CoV-2 in HCWs. Here, as reported for adolescents receiving two vaccines,[18,19] post-V1, infection-naive adolescents generated higher magnitude anti-S IgG responses than infection-naive adults (49,696 vs 33,339, x1.5, p=0.03, Mann-Whitney test) and previously-infected adolescents generated greater anti-S IgG responses than previously-infected adults (743,691 vs 269,985, x2.9, p<0.0001, Mann-Whitney test) (Fig. 4A). Post-V1 nAb responses did not differ significantly between adolescents and adults (Fig. 4B).

**Figure 4:**
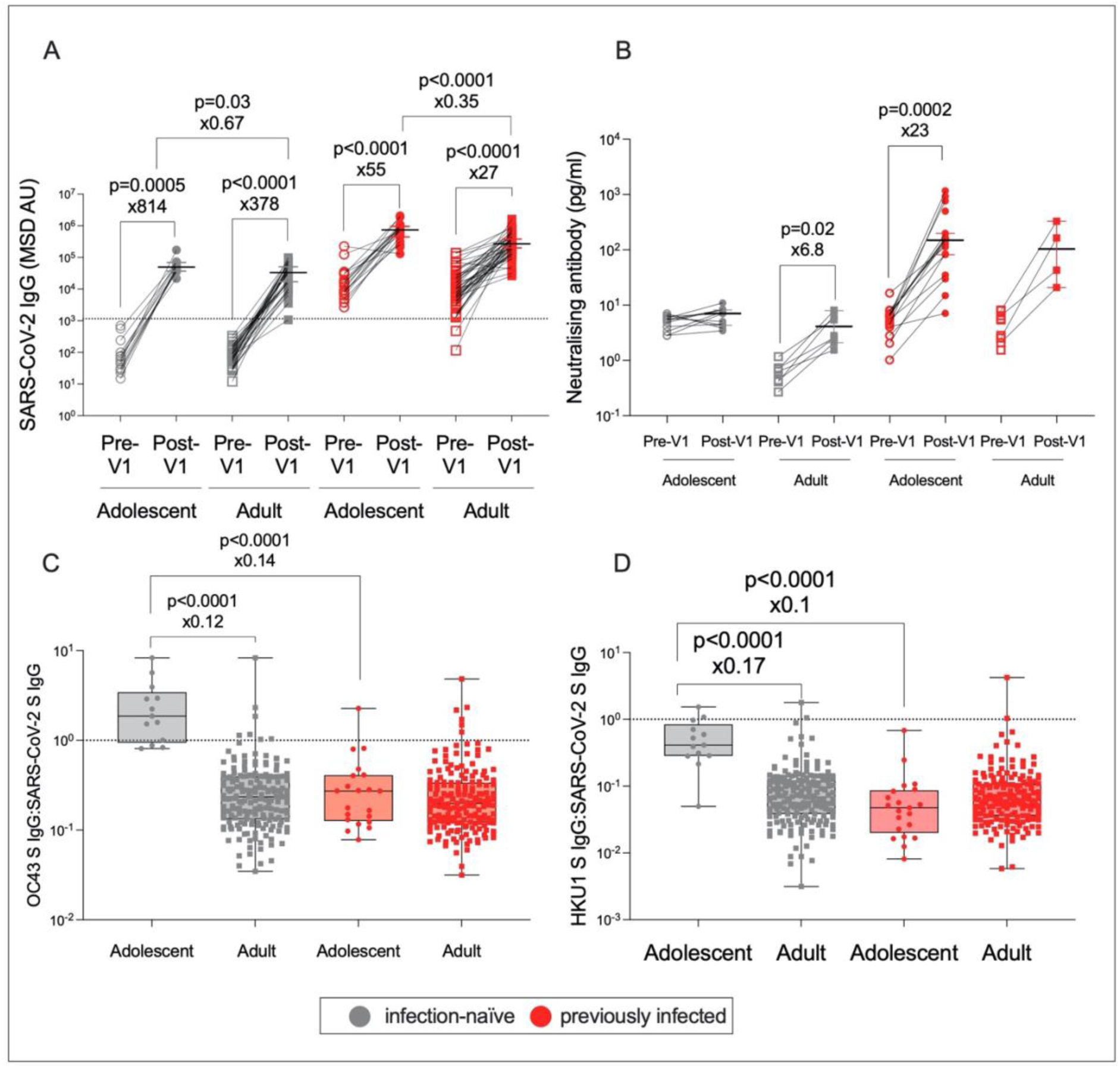
Age-specific effects in the humoral response to BNT162b2. IgG targeting S in infection-naïve adolescents (grey circles), infection-naïve adults (32-52 years) (grey squares), previously-infected adolescents (red circles) and previously-infected adults (red squares), pre-V1 (unfilled shapes) and post-V1 (filled shapes) as measured by an MSD v-plex immunoassay (**A**). nAb concentration targeting S in infection-naïve and previously-infected adolescents and adults as measured by an MSD ACE2-Spike binding immunoassay (**B**). The ratio of IgG targeting HCoV-OC43 S to SARS-CoV-2 S (C) and the ratio of IgG targeting HCoV-HKU1 S to SARS-CoV-2 S (D) in infection-naive adolescents (grey circles), infection-naive adults (grey squares), previously-infected adolescents (red circles), and previously-infected adults (red squares). P-values represent Mann-Whitney test values for unpaired data, and Wilcoxon signed-rank test values for paired data. Fold change calculated as the ratio of population medians.

To investigate why infection-naive adolescents generate relatively weak nAb responses post-V1, despite a strong total IgG response, we sought to address the hypothesis that cross-reactive antibody responses to endemic HCoVs might be present at higher levels in infection-naïve adolescents, thereby interfering with the generation of novel SARS-CoV-2-specific responses to BNT162b2, as has been suggested previously.[28,29] Humoral responses to HCoVs have been associated with worse COVID-19 outcomes, through the inhibition of novel responses to SARS-CoV-2 as a result of immune imprinting or ‘original antigenic sin’.[28] Pre-existing cross-reactive IgG may promote higher magnitude IgG responses to the conserved S2 region of SARS-CoV-2 S following vaccination, whilst it is the less conserved S1 region that is the target of most neutralising antibodies.[30] Children have been reported to display higher immunity to endemic HCoVs than adults,[31] perhaps due to high circulation of viruses in schools.

Our data supported the hypothesis that cross-reactive antibody responses to HCoVs are associated with weaker vaccine-induced neutralising responses: in this study, the ratio of IgG targeting betacoronaviruses HCoV-HKU1 and HCoV-OC43 S to IgG targeting SARS-CoV-2 S was significantly higher in infection-naive adolescents versus infection-naive adults (0.4 vs 0.06, x5.9, p<0.0001; 1.9 vs 0.2, x8.3, p<0.0001, Mann-Whitney tests) and versus previously-infected adolescents (0.4 vs 0.04, x10, p<0.0001; 1.9 vs 0.3, x7.1, p<0.0001, Mann-Whitney test) post-V1 (Fig. 4CD). Furthermore, the ratio of IgG targeting HCoV-HKU1 and HCoV-OC43 S to IgG targeting SARS-CoV-2 S was significantly negatively correlated with nAb response (HKU1: r=-0.75, p<0.0001; OC43: r=-0.84, p<0.0001) in all adolescents, though this significance was lost when adolescents were divided into infection-naïve and previously-infected.

### Sex differences in response to BNT162b2 vaccination

Females typically make stronger IgG responses than males following vaccination,[10,11,32,33] including after influenza vaccines.[9,16] Surprisingly, therefore, here, infection-naïve males generated significantly higher post-V1 IgG targeting both SARS-CoV-2 S and RBD than females (62,270 vs 36,951, x2, p=0.008; 23,860 vs 11,443, x2, p=0.02, respectively, Mann-Whitney tests) (Fig. 5AB). There was no significant difference in IgG response between the sexes in previously-infected individuals (Fig. 5CD). Furthermore, there was a trend towards a stronger RBD and S nAb response post-V1 in infection-naive males compared to infection-naive females, although this was not significant (p=0.07 and p=0.15, respectively, Mann-Whitney tests) (Fig. 5EF). There was no significant different in baseline IgG responses between males and females. There was no sex difference in the humoral response to BNT162b2 in adults.[26]

**Figure 5:**
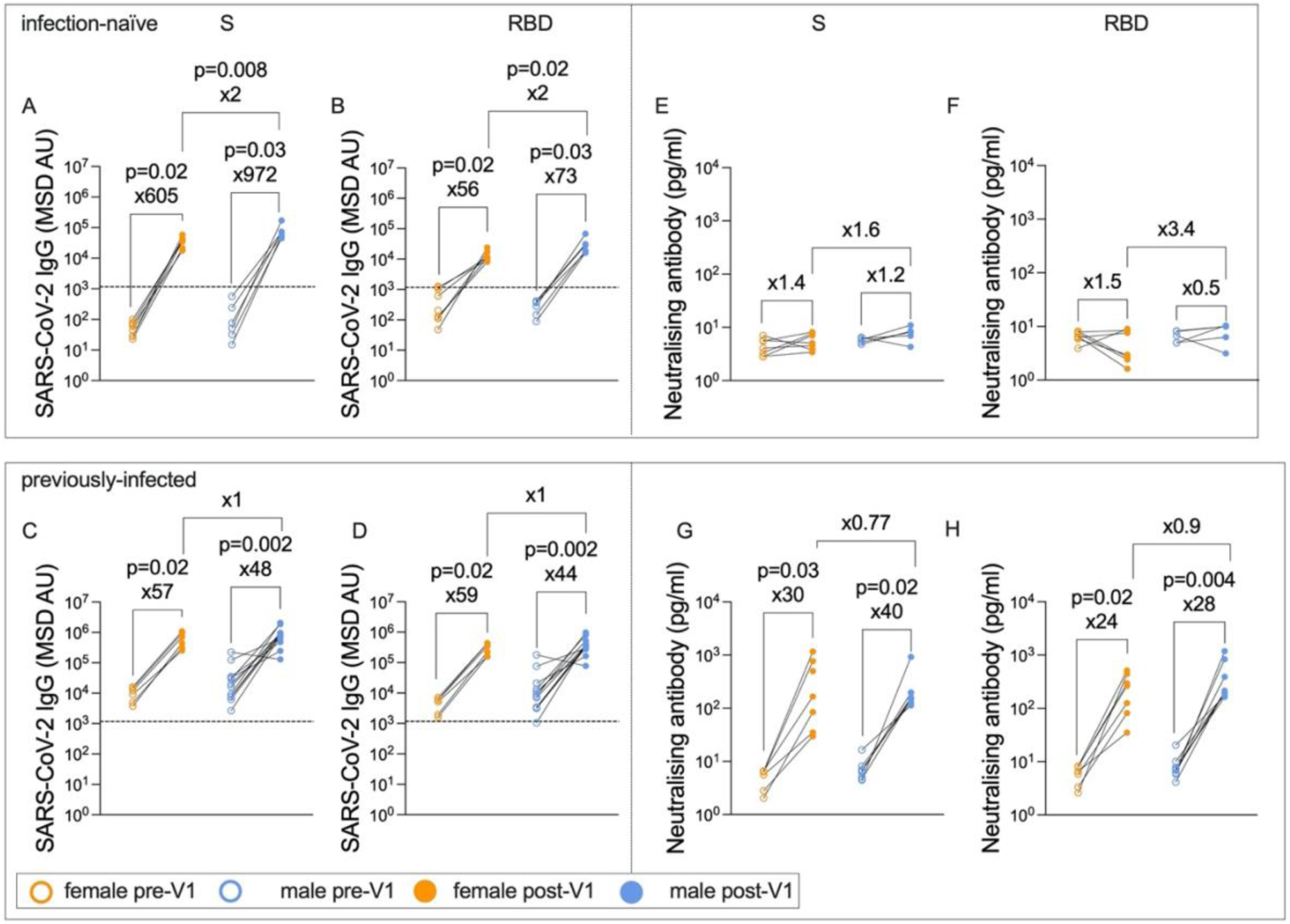
Infection-naïve males generate greater post-V1 IgG responses than females. IgG targeting S (**A**) and RBD (**B**) in infection-naive adolescents pre-V1 (unfilled circles) and post-V1 (filled circles) in females (orange circles) and males (blue circles) as measured by an MSD v-plex immunoassay. IgG targeting S (C) and RBD (D) in previously-infected adolescents pre-V1 and post-V1 in females and males. Concentration of nAbs targeting S (E) and RBD (F) in infection-naive adolescents as measured by an MSD ACE2-Spike binding inhibition assay. Concentration of nAbs targeting S (G) and RBD (H) in previously-infected adolescents. P-values represent Wilcoxon test values for paired data and Mann-Whitney test values for unpaired data. Fold change calculated as the ratio of population medians.

A potential mediator of immune sex differences is the effect of sex hormones. Males enter puberty on average 2 years later than females. To ensure males in this cohort had entered puberty, steroid hormones including testosterone, dihydrotestosterone (DHT) and progesterone were measured by Tandem mass spectrometry; these data are the focus of a future publication. All but the two youngest males (12 years 2 months and 12 years 10 months) demonstrated pubertal androgen levels. Testosterone correlated with age in males only (r=0.47, p=0.05).

### Humoral responses to LAIV administration

As well as the immune response to BNT162b2, co-administration of the LAIV enabled the characterisation of immunity against influenza following vaccination. To determine the effect of the LAIV on lineage-specific anti-haemagglutinin (HA) IgG titres, enzyme-linked immunosorbent assays (ELISAs) were performed on pre- and post-LAIV samples for the 26 individuals who received the LAIV (Fig. 6). As expected, IgG titres were significantly higher post-LAIV for A/Cambodia (H3N2), A/Victoria (H1N1), and B/Phuket (Yamagata) (9.3 vs 13.9, x1.5, p<0.0001; 11 vs 13.4, x1.2, p=0.0002; 7 vs 10.2, x1.5, p<0.0001; respectively, Wilcoxon signed-rank tests) (Fig. 6A). Surprisingly, post-LAIV anti-HA IgG responses towards the B/Washington (Victoria) lineage were not significantly increased compared to pre-LAIV. A possible explanation lies in the observation that responses to B/Washington (Victoria) were strongly correlated with age for both pre- and post-vaccine timepoints (r=0.61, p=0.0001, r=0.57, p=0.0008, respectively, Spearman rank test) (Fig. 6BC). By contrast, there was no correlation between age and post-LAIV IgG for A/Cambodia (H3N2) or B/Phuket (Yamagata), and for A/Victoria (H1N1) pre-LAIV IgG levels only were weakly correlated with age (r=0.39, p=0.02, Spearman rank test). This pattern suggests that natural exposure to B/Washington (Victoria) is so frequent in this cohort that vaccination against this strain of influenza does not add significantly to the natural immunity that is accumulated over adolescence. Pre-existing immunity to influenza has been widely described, from prior infection and vaccination, in support of this finding. [34,35]

**Figure 6:**
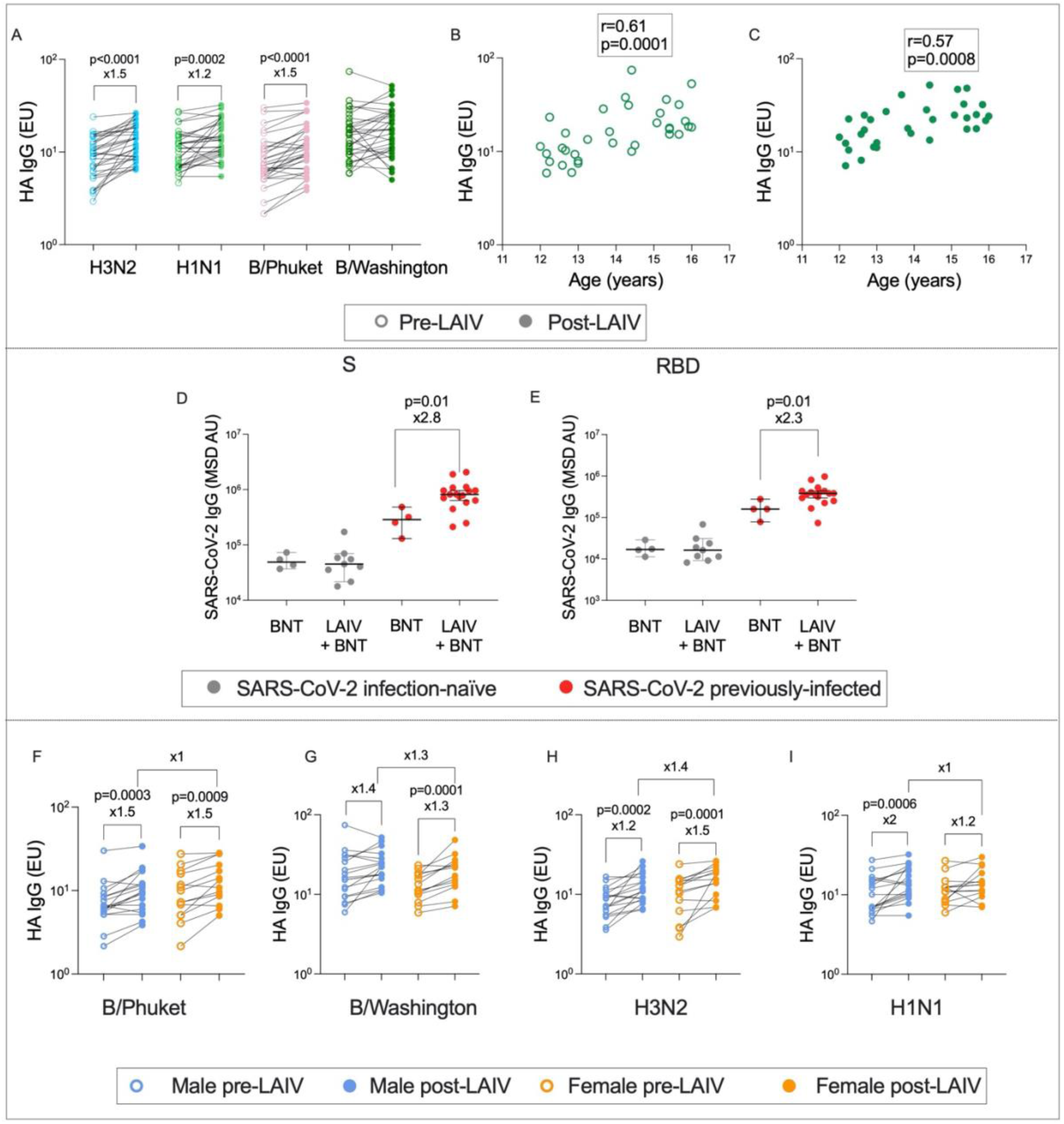
Age- and sex-specific immunity to influenza following LAIV administration. IgG targeting haemagglutinin (HA) pre- (unfilled circles) and post- (filled circles) LAIV administration for the four influenza lineages (**A**) (P-values from Wilcoxon tests). The correlation between age and IgG targeting HA for the B/Washington lineage pre-LAIV (**B**) and post-LAIV (**C**) (Spearman rank r- and p-values). IgG targeting SARS-CoV-2 S (**D**) and RBD (**E**) in infection-naïve (grey) and previously-infected (red) adolescents who received the BNT162b2 vaccine alone (BNT), or co-administered with the LAIV (LAIV + BNT) (Mann-Whitney p-values). IgG targeting HA pre- (unfilled circles) and post- (filled cirles) LAIV administration in males (blue) and females (orange) for the four influenza lineages (**F-I**) (Wilcoxon p-values).

Unexpectedly, and in contrast to previous studies,[36] adolescents previously-infected with SARS-CoV-2 who received both BNT162b2 and the LAIV generated significantly higher post-V1 IgG targeting both S and RBD compared to adolescents previously-infected with SARS-CoV-2 who received BNT162b2 alone (286,185 vs 817,284, x2.8, p=0.01; 159,101 vs 379,429, x2.3, p=0.01, Mann-Whitney tests), although this analysis involved very small numbers of individuals (Fig. 6DE). We did not find a sex difference in the IgG response to the LAIV (Fig 6F-I).

## Discussion

Understanding the quantitative markers of vaccine immunogenicity, as well as confounding patient demographic factors, will help better define correlates of protection against SARS-CoV-2 and improve interpretability of future vaccine trials. Due to the discrepancy between IgG and nAb response in infection-naive adolescents, these data support the use of nAb titre as well as total IgG when assessing vaccine immunogenicity.[37,38] Other studies have established that BNT162b2 and the CoronaVac inactivated virus vaccine elicit robust nAb responses post-V2 in infection-naive adolescents.[19,39] The totality of data described herein suggest that a robust nAb response is prompted in infection-naive adolescents after two doses but previously-infected adolescents only require one dose. Previous studies in adults have differed in their evaluation of vaccine-induced versus infection-induced humoral immunity, but these data show that a similar IgG response is elicited in adolescents after natural infection and one vaccine dose compared to two vaccine doses only.[40] The longevity of these responses is uncertain due to the lack of an extended follow up in this cohort but should be the focus of future studies.

Other research has shown that two doses of BNT162b2 elicit robust T_H_1 T-cell responses in adults, with widespread interferon-gamma (IFNγ) production.[26,41] S-specific T-cell responses following vaccination with BNT162b2 were generated post-V2, but not post-V1, in another cohort of infection-naive adolescents.[18] This contrasts with the data described herein, where one dose of BNT162b2 is sufficient to induce a significant increase in S-specific CD4+ T-cells in infection-naive adolescents. The lack of a significant reduction in T-cell response across timepoints suggests that these responses are stable over several months, as reported elsewhere.[42,43] Similarly to the IgG response, SARS-CoV-2-specific T-cell responses post-V1 in previously-infected individuals reach similar frequencies to post-V2 in infection-naive individuals.

BNT162b2 has been shown to promote greater IgG production in adolescents compared to adults post-V1.[18] Similarly, here, both infection-naive and previously-infected adolescents generated stronger IgG responses than adults. This may result from higher exposure to endemic HCoVs in adolescents, promoting a stronger IgG response to conserved SARS-CoV-2 antigens. However, only previously-infected adolescents generated a strong and broad nAb response targeting multiple variants, and infection-naïve adolescents appeared to rely more on cross-reactive antibodies following their first dose of BNT162b2 compared to both infection-naïve adults and previously-infected adolescents. One interpretation for these patterns is immune imprinting, wherein prior exposure to circulating endemic coronaviruses negatively impacts vaccine-induced immunity. Higher levels of cross-reactive IgG have been described in children compared to adults,[31,44,45] and this may result in a stronger memory B-cell response that is weakly neutralising following the first dose of BNT162b2 in infection-naive adolescents. In previously-infected adolescents, prior exposure to SARS-CoV-2 may overcome immune imprinting and enable a robust nAb response. This is supported by the strong negative correlation between HCoV:SARS-CoV-2 IgG ratio and nAb response.

Immune responses to many adult and childhood vaccines, as well as responses to natural infection with viral pathogens, are consistently higher in females and associated with increased inflammation and autoimmunity as well as CD4+-skewed T-cell responses and greater B cell activation and IgG production. [10,11,32,33] Female IgG responses to influenza vaccines have been shown to reach twice the magnitude of male IgG responses, and females also report more frequent SAEs to viral vaccines.[11,33] One exception to this trend is COVID-19 mRNA vaccines, for which vaccine-induced myocarditis is more frequent in young males. [13,14,46] Notably, in this cohort, we observe increased post-V1 anti-S and anti-RBD IgG responses in infection-naive males compared to infection-naive females, in contrast to expectations based on other vaccines such as inactivated influenza vaccines.[9,11] We did not observe a significant sex difference in anti-HA IgG titres following the LAIV - this is surprising in the context of established literature,[10,11] but may be obscured by the very small increase in anti-HA IgG post-V1 in this cohort, the effect of a live-attenuated rather than inactivated influenza vaccine, the use of different serological assays, or the result of co-administration with BNT162b2.[47] Furthermore, there was no sex difference in anti-SARS-CoV-2 IgG for either infection-naïve or previously-infected adults from the PITCH dataset.

A potential confounder for this study is that adolescents of this age group are likely at different stages of puberty and therefore have diverse levels of testosterone, oestrogen and progesterone. Furthermore, males experience puberty at older ages compared to females, and therefore the sex difference identified herein may result from the confounding effects of puberty. If many male adolescents had not gone through puberty at the time of sampling, the increased humoral responses to vaccination in males may result from an absence of the immunosuppressive effects of androgens. Steroid hormones were measured in this cohort, and although these results are the focus of a future publication, it was identified that all but two males had pubertal levels of testosterone. This promotes confidence in the results of comparisons between sexes, as the majority of males had undergone puberty at time of sampling.

Finally, the correlation between B/Washington influenza IgG responses with age in 12–16-year-olds, as well as the lack of anti-B/Washington HA IgG boosting following the LAIV, suggests recent exposure to the B/Washington strain of influenza in this cohort. Our findings that co-administration of BNT162b2 with the LAIV improves IgG response in previously-infected individuals is in contrast with findings for NVX-COV2373, where co-administration with inactivated quadrivalent influenza vaccines reduced SARS-CoV-2-specific IgG titres[36]. However, studies of co-administration of COVID-19 mRNA vaccines with quadrivalent influenza vaccines in adults have reported no reduction in antibody response compared to administration of mRNA vaccines alone.[48,49] A potential explanation for improved anti-S IgG responses following co-administration may be increased innate immune activation due to intranasal LAIV administration, particularly in the nasal mucosa, leading to greater SARS-CoV-2-specific local T-helper cell activation.

This study has some limitations. The small numbers of adolescents assayed in this cohort make broad conclusions difficult, particularly when making comparisons between small sub-groups such as co-administered LAIV/BNT162b2 and BNT162b2-alone individuals. No mucosal samples were taken and so mucosal immunity is not assessed in this cohort. Neutralisation responses to SARS-CoV-2 were estimated using the MSD-ACE2 inhibition assay. This has shown to correlate with live virus assays,[20,21,24] but live virus neutralisation is likely a more accurate measure of nAbs. In addition, no neutralisation assays were carried out for influenza lineages, which would have shed further light on the functionality of humoral immunity to influenza. Furthermore, the lack of an extended follow-up in this study makes assessments of immune durability impossible but should be the focus of future studies.

Taken together, these data paint a complex picture of vaccine-induced immunity in adolescents, with a potential role for immune sex and age differences in determining antibody responses to vaccination. These findings have important implications for paediatric vaccination regimes, such as the potential benefit of co-administration with influenza vaccines, and the necessity to consider sex and age when studying vaccine-induced immunity.

## Materials and Methods

### Ethics

This longitudinal cohort study was conducted from November 2021 to February 2022. Eligible participants were healthy adolescents aged 12-16 were who either had no history of SARS-CoV-2 infection or had experienced mild disease prior to enrolment. Eligible participants were identified via their participation in school-based vaccination events. Written informed consent was obtained from all patients and ethical approval was given by the Central University Research Ethics Committee (reference: CUREC R71346/RE001). 32–52-year-old healthy HCWs were recruited as part of the PITCH consortium of HCWs under the GI Biobank Study 16/YH/0247, approved by the research ethics committee (REC) at Yorkshire & The Humber - Sheffield Research Ethics Committee on 29 July 2016, which has been amended for this purpose on 8 June 2020.

### Sample collection and processing

For the BNT162b2 vaccination (dose 1 (V1) and dose 2 (V2)), patients received 30 ug of vaccine intramuscularly. LAIV was administered immediately after V1 only; patients received 0.1mL intranasally in both nostrils. Whole blood samples from all 34 individuals were taken immediately before V1 (sample pre-V1). Samples from all 34 individuals were taken a mean of 37 days after V1 (33-39]) (sample post-V1), from 23 individuals 2 days before V2 (0-8) and 96 days after V1 (81-114) (sample pre-V2), and from 14 individuals 35 days after V2 (30-40) (sample post-V2). All whole blood samples were processed the same day as collection as described in the methods. All serum samples were tested for anti-Spike (S) and anti-nucleocapsid (N) IgG; individuals were classified as seropositive if their anti-N IgG titre was above the previously determined MSD immunoassay cut-off at any point in the study or if their anti-spike (S) IgG titre was above the cut-off pre-V1[26]. The percentage of seropositive patients increased from 52% to 71% over the course of the study.

Whole blood samples were transported from their collection site to an academic laboratory and processed the same day. PBMCs and plasma were isolated as described elsewhere[51]. Briefly, PBMCs were isolated using Lymphoprep (1.077 g/ml, Stem Cell Technologies) through density gradient centrifugation. Plasma and PBMCs were collected and plasma was spun at 2000g for 10 minutes to remove platelets. PBMCs were washed twice with RPMI 1640 (Sigma, USA) containing 10% heat inactivated foetal calf serum, 2mM L-Glutamine and 1mM penicillin/streptomycin (Sigma). An estimated ten million cells were resuspended in media and counted using a Muse Cell Analyser (Luminex Corporation, USA). Plasma and PBMCs were frozen and stored at 80°C for later use. Steroid hormone concentrations were quantified by tandem mass spectrometry by collaborators at Imperial College London.

### MSD serological assays

IgG responses to SARS-CoV-2 S, N and RBD as well as the S proteins of HCoV-OC43, HCoV-NL63, HCoV-229E, HCoV-HKU1, SARS-CoV-1 and MERS-CoV were measured using a Meso Scale Diagnostics (MSD) V-plex immunoassay ‘Coronavirus panel 3’ (MSD, USA) according to the manufacturer’s protocol. Plates were incubated in Blocker A solution for 30 minutes at room temperature (RT) with shaking at 700rpm. Plasma or serum was diluted at 1:1000 and 1:10000 in Diluent 100, and a seven-point standard curve of MSD reference standard beginning at 1:10 was prepared in duplicate. Three internal controls and an in-house control of convalescent serum were also used, with Diluent 100 used as a blank. Plates were washed three times with MSD Wash Buffer and samples and standards added to the plate before incubation at RT for two hours with shaking at 700rpm. Plates were washed three times with MSD Wash Buffer and detection antibody solution was added. Plates were incubated for one hour at RT with shaking at 700rpm. Plates were washed three times with MSD Wash Buffer. Neat MSD Gold Read Buffer was added, and plates were read immediately on a MESO QuickPlex SQ 120 (MSD, USA). Data was analysed using MSD Discovery Workbench software. Thresholds for seropositivity were taken from analyses of pre-pandemic sera, as published elsewhere[26], and defined as 1160 AU/ml for SARS-CoV-2 S, 1169 for RBD, and 3874 for N.

nAb titres were quantified using Meso Scale Diagnostics ACE2 inhibition assays, ‘Panel 27’, (analytes: SARS-CoV-2 S, SARS-CoV-2 S (B.1.351), SARS-CoV-2 S (B.1.617.2; AY.4), SARS-CoV-2 S (BA.2), SARS-CoV-2 S (BA.2.12.1), SARS-CoV-2 S (BA.2+L452M), SARS-CoV-2 S (BA.2+L452R), SARS-CoV-2 S (BA.3), SARS-CoV-2 S (BA.4), SARS-CoV-2 S (BA.5)) according to the manufacturer’s instructions. Plates were incubated in Blocker A solution for 30 minutes at RT with shaking at 700rpm. Serum was diluted at 1:10 and 1:100, and a seven-point standard curve of MSD calibration reagent was prepared with 4-fold serial dilutions. Plates were washed three times with MSD Wash Buffer and samples and calibrator were added to the plate. Plates were incubated at RT for one hour with shaking at 700rpm. Sulfo tagged ACE2 protein was added to the plate and incubated at RT for one hour with shaking at 700rpm. Plates were washed three times with MSD Wash Buffer and MSD Gold Read Buffer was added. Plates were read immediately on a MESO QuickPlex SQ 120 (MSD, USA). Data was analysed using MSD Discovery Workbench software.

### Influenza ELISA assay

IgG responses to influenza A/Victoria (H1N1), B/Washington (Victoria), A/Cambodia (H3N2), and B/Phuket (Yamagata) HA antigens were measured using an indirect ELISA. HA antigens (The Native Antigen Company, Oxford) were diluted to 1ug/ml in PBS and used to coat 535 Nunc-Immuno 96-well plates (Thermo Fisher Scientific, USA) overnight at 4°C (A/Victoria/2570/2019 (H1N1)pdm09-like virus (NCBI Accession Number: EPI1799581), amino acids 1-528 and C-terminal His-tag; Cambodia/e0826360/2020 (H3N2)-like virus (NCBI Accession Number: EPI1799580), amino acids 46-469 and C-terminal His-tag; B/Washington/02/2019 (B/Victoria lineage)-like virus (NCBI Accession Number: EPI1846769), amino acids 31-469 and C-terminal His-tag, B/Phuket/3073/2013 (B/Yamagata Lineage)-Like virus] (NCBI Accession Number: EPI1799823), amino acids 44-466 and C-terminal His-tag) Plates were washed three times in 0.1% PBS-Tween, before blocking with Casein-PBS Buffer for one hour at RT. Plasma was diluted 1:200 in Casein-PBS Buffer and added to plates in duplicate. A ten-point standard curve of pooled highly reactive sera beginning at 1:25 was prepared in duplicate and added to plates. Casein-PBS Buffer was used as a negative control. Plates were incubated for two hours at RT and washed six times in 0.1% PBS-Tween. Secondary antibody – goat anti-human IgG conjugated to alkaline phosphatase (Sigma, USA) was diluted 1:1000 in Casein-PBS Buffer and added to plates. Plates were incubated for one hour at RT before washing six times in 0.1% PBS-Tween. 4-nitrophenyl phosphate in diethanolamine buffer (Pierce, Loughborough, UK) was added as a substrate and plates were incubated for 15 minutes. 405nm absorbance was read using an ELx800 microplate reader (Cole Parmer, London, UK).

### Proliferation assay

T-cell responses were assayed using a CellTrace Violet Proliferation assay as described elsewhere[51]. Cryopreserved PBMCs were thawed in 30mL RPMI containing 10% human AB serum (Sigma), 2mM L-Glut and 1mM Pen-Strep. Cells were washed twice with PBS and stained with CellTrace Violet (Life Technologies) at 2.5uM for 10 minutes at RT. Cold FCS was added to quench the reaction. Cells were plated at 250,000 cells per well in a 96-well round-bottom plate. Peptide pools covering SARS-CoV-2 S1, S2, M and N, as well as HCoV-OC43 and HCoV-HKU1 S, were added to stimulate cells at a final concentration of 1ug/ml (Mimotopes, USA) (Supplementary File 1). Media containing 0.1% DMSO (Sigma) was used as a negative control. Phytohaemagglutinin L (Sigma) was used as a positive control at a final concentration of 2 ug/ml. Plates were incubated at 37°C, 5% CO2, 95% humidity for 7 days, with a hemimedia change at day 4. On day 7, cells were washed in PBS and stained with fluorochrome-conjugated antibodies for CD4, CD8, and CD3 in PBS. LIVE/DEAD Fixable Aqua was used as a viability marker (Thermo Fisher). Cells were fixed in 4% paraformaldehyde (Sigma) for ten minutes at 4°C and washed in PBS before storing at 4°C in the dark before being run on an MACSquant X (Miltenyi). Gating strategy can be viewed in Supp. Fig. 8.

### Statistical analysis

All analyses were performed in GraphPad Prism 9.0. For pairwise comparisons, two-tailed Mann-Whitney tests were used for unpaired data and Wilcoxon signed-rank tests for paired data. For correlations, Spearman rank tests were used.

## Contributions

CJ: Conceptualisation, Formal analysis, Investigation, Writing – Original Draft Preparation, Review & Editing, Visualisation EA: Investigation, Data Curation, Writing – Review & Editing, Project administration AC: Investigation, Data Curation, Writing – Review & Editing, Project administration NL: Investigation, Writing – Review & Editing SL: Formal analysis, Investigation, Writing – Review & Editing AO: Investigation, Writing – Review & Editing JR: Writing - Review & Editing OS: Investigation, Writing – Review & Editing PITCH Consortium: Investigation, Formal analysis, Data Curation CT: Resources, Writing – Review & Editing, Methodology EB: Conceptualization, Writing – Review & Editing, SD: Supervision, Writing – Review & Editing LT: Supervision PK: Supervision, Writing – Review & Editing, Conceptualization, Methodology MC: Supervision, Writing – Review & Editing, Methodology PG: Conceptualization, Methodology, Resources, Writing – Review & Editing, Supervision, Project administration, Funding acquisition

## Funding

P.K. is is funded by WT109965MA and is an NIHR Senior Investigator. S.J.D. is funded by an NIHR Global Research Professorship (NIHR300791). E.B. Is funded by an NIHR Senior Investigator award; the views expressed do not represent those of the NIHR or the NHS. The PITCH consortium was funded by the UK Department of Health and Social Care and UKRI (MR/W02067X/1), with contributions from UKRI/NIHR through the UK Coronavirus Immunology Consortium (MR/V028448/1), the Huo Family Foundation and The National Institute for Health Research (UKRIDHSC COVID-19 Rapid Response Rolling Call, Grant Reference Number COV19-RECPLAS).

## Data Availability

All data produced in the present study are available upon reasonable request to the authors

## Acknowledgements

We are grateful to staff at the schools from which the participants were enrolled and to the participants and their families.

The authors declare no competing interests.

## Notes

### Competing Interest Statement

The authors have declared no competing interest.

### Author Declarations

The Central University Research Ethics Committee of University of Oxford gave ethical approval for this work.

