## Supplementary Figures for "Age- and sex-specific differences in immune responses to BNT162b2 COVID-19 and live-attenuated influenza vaccines in UK adolescents"

**Supplementary Figure 1: Fold change in nAb and IgG titre in adolescents post-V1 and post-V2**

Fold change in nAbs targeting S (**A**) and RBD (**B**) in infection-naive adolescents (grey circles), and previously-infected adolescents (red circles) post-V1 and post-V2 as measured by an MSD ACE2-S binding immunoassay. Fold change in IgG targeting S (**D**) and RBD (**D**) in infection-naive and previously-infected adolescents as measured by an MSD v-plex immunoassay. P-values represent Mann-Whitney test values.

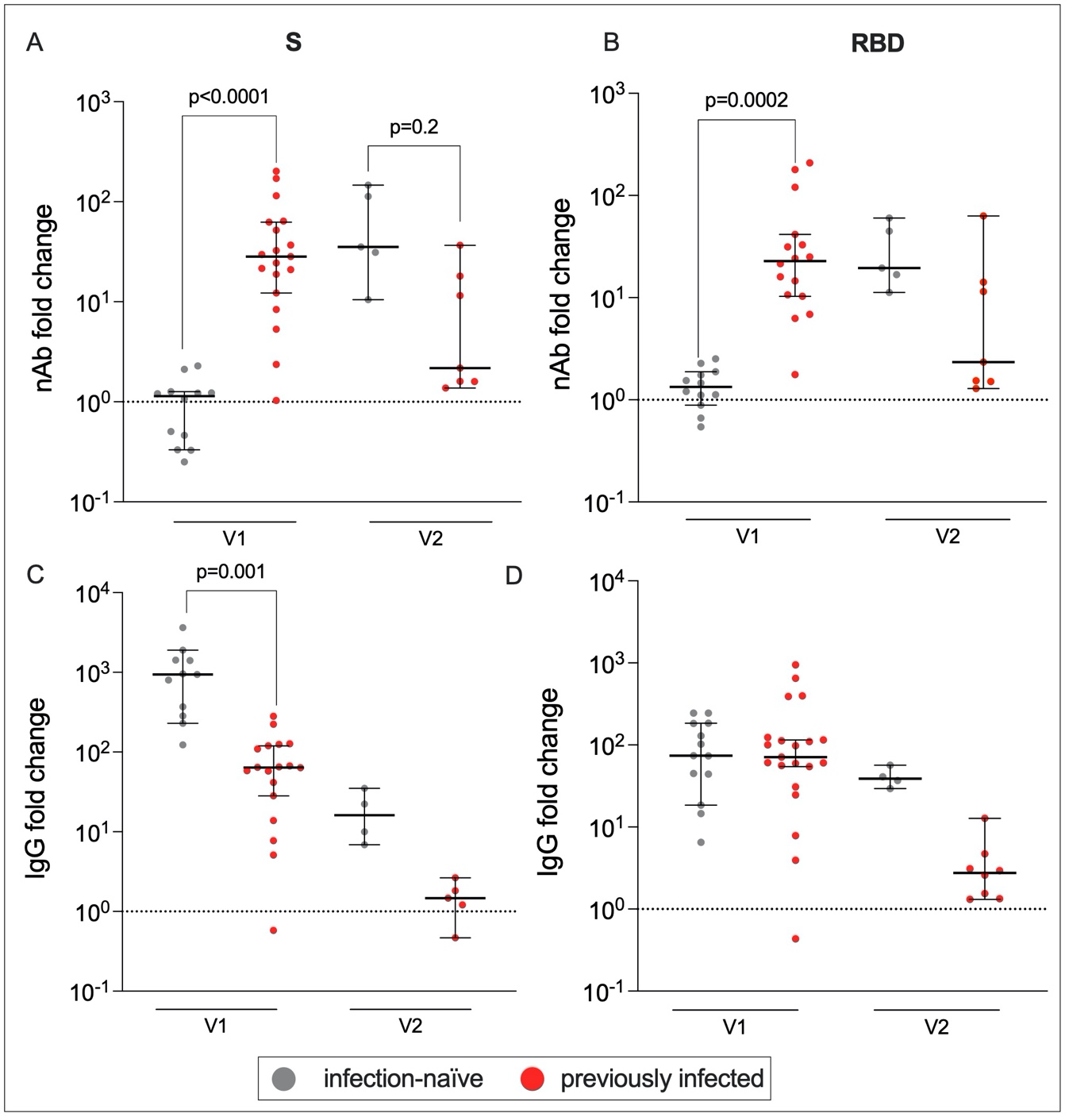

**Supplementary Figure 2: nAb responses to SARS-CoV-2 variants.**

Percent inhibition of ACE2-S binding for common variants in infection-naïve (grey) and previously-infected (red) adolescents. P-values from Wilcoxon tests.
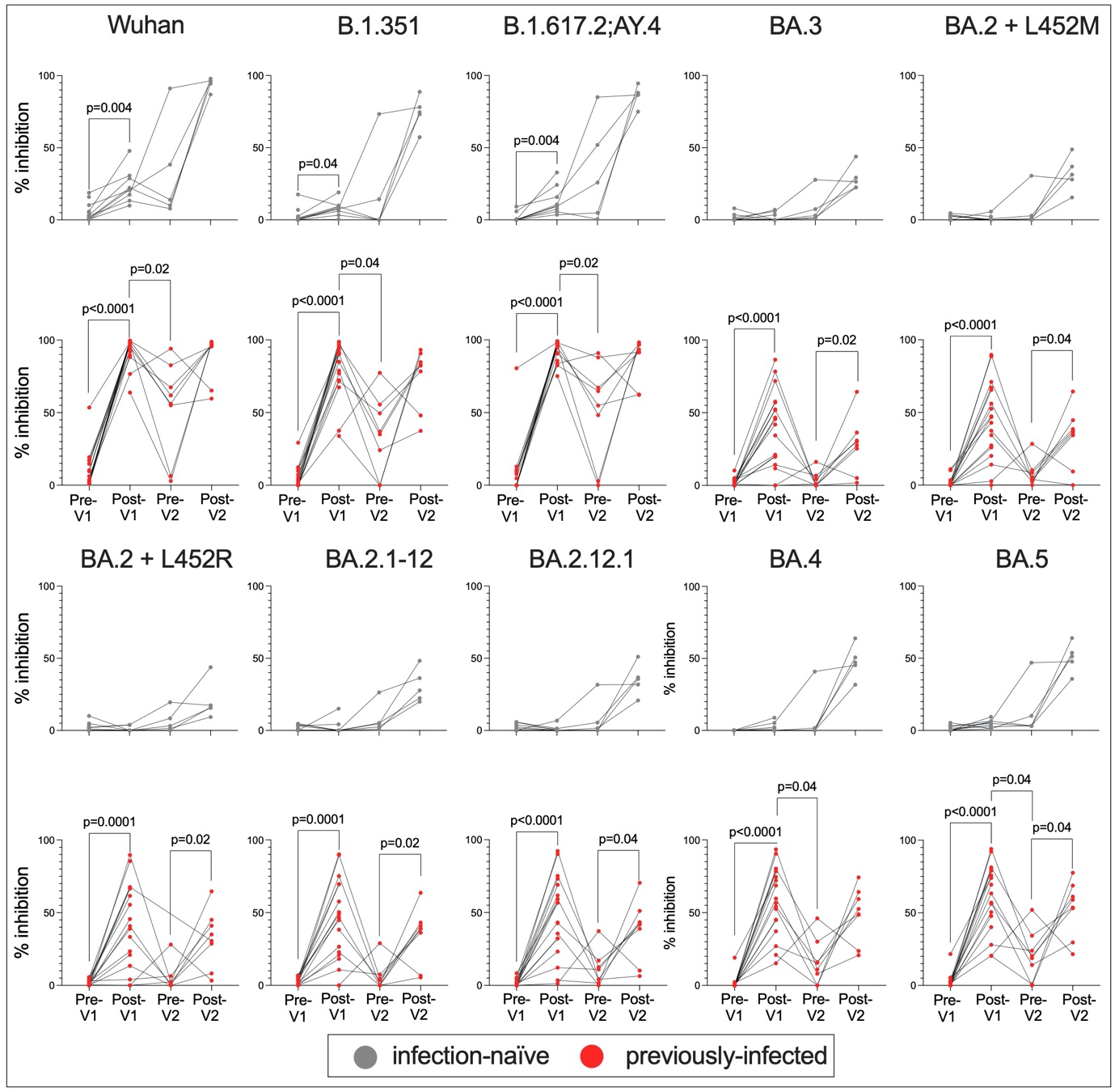

**Supplementary Figure 3: Cellular responses to endemic HCoVs.** Proliferating CD4+ and CD8+ T-cells targeting HCoV-OC43 S2 (A) and HCoV-HKU1 S2 (B) in infection-naïve (grey) individuals. % proliferating CD4+ and CD8+ T-cells targeting HCoV-OC43 S2 (C) and HCoV-HKU1 S2 (D) in previously-infected individuals (red). Values below 1% were given nominal values of 0.9%.

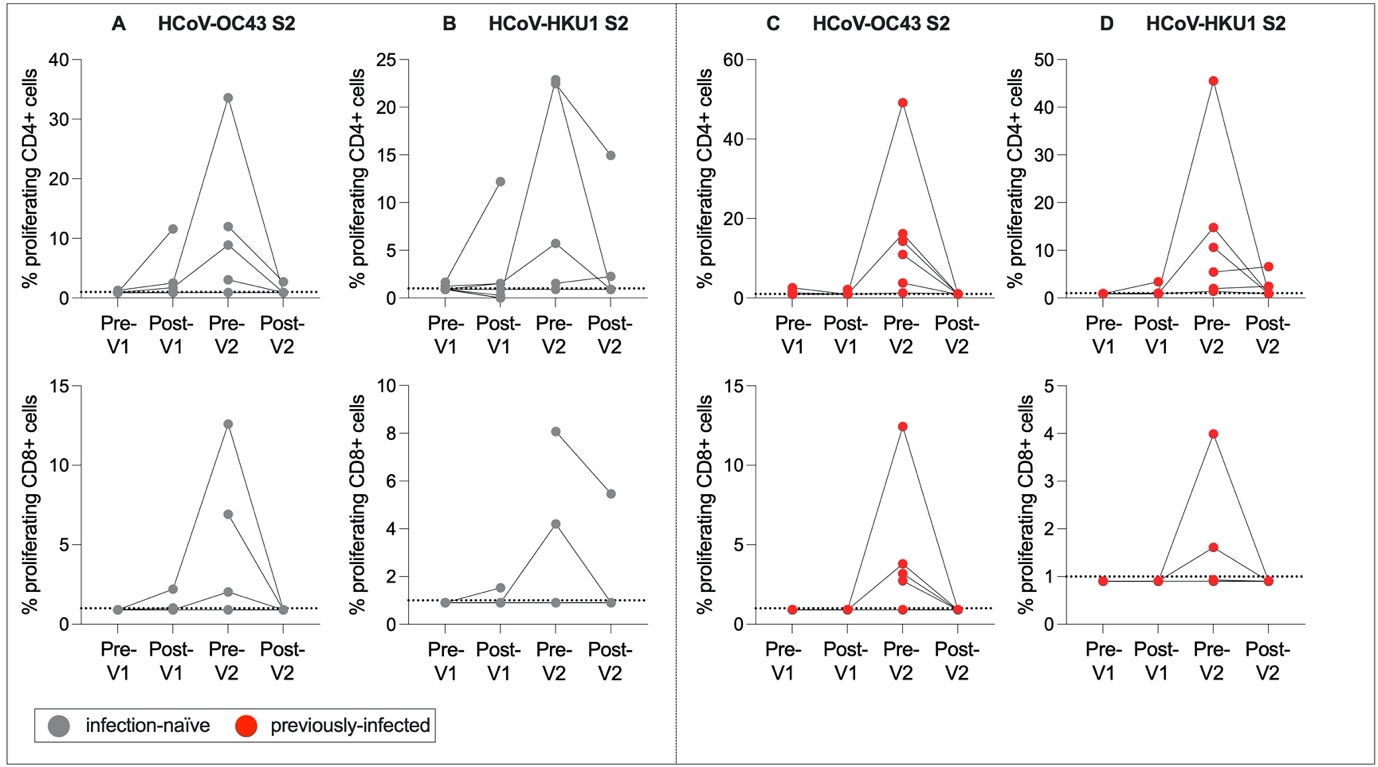

**Supplementary Figure 4: Gating strategy for T-cell proliferation assay.**

Gates were drawn on lymphocytes (SSC-A x FSC-A), single cells (FSC-H x FSC-A), live cells (CD3 x Live/Dead stain), and CD4+ and CD8+ cells (CD4 x CD8). Proliferating CD4+ cells were gated as compared to a negative control (CD4 x CTV). Proliferating CD8+ cells were gated as compared to a negative control (CD8 x CTV).
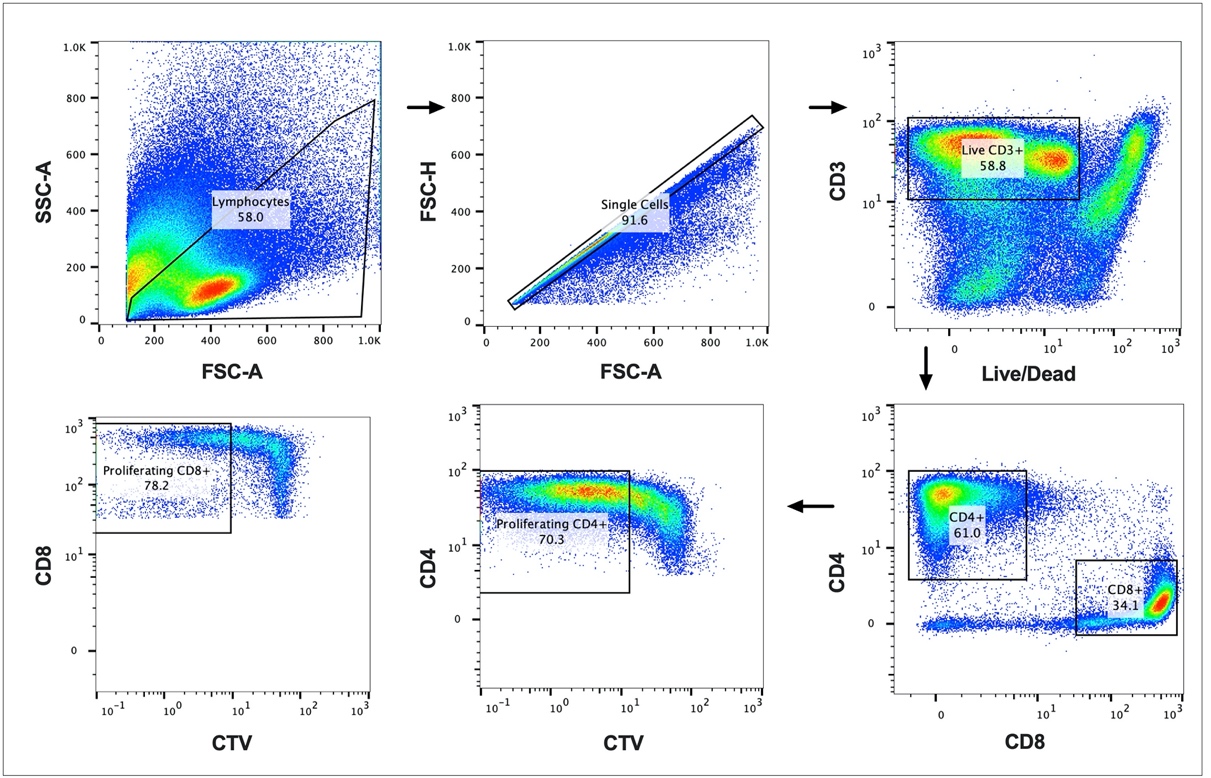
